# Equivalency of Protection from Natural Immunity in COVID-19 Recovered Versus Fully Vaccinated Persons: A Systematic Review and Pooled Analysis

**DOI:** 10.1101/2021.09.12.21263461

**Authors:** Mahesh B. Shenai, Ralph Rahme, Hooman Noorchashm

## Abstract

**BACKGROUND:** We present a systematic review and pooled analysis of clinical studies to date, that (1) specifically compare the protection of natural immunity in the COVID-recovered versus the efficacy of full vaccination in the COVID-naive, and (2) the added benefit of vaccination in the COVID-recovered, for prevention of subsequent SARS-CoV-2 infection.

**METHODS:** Using the PRISMA 2020 guidance, we first conducted a systematic review of available literature on PubMed, MedRxIV and FDA briefings to identify clinical studies either comparing COVID vaccination to natural immunity or delineating the benefit of vaccination in recovered individuals. After assessing for eligibility, studies were qualitatively appraised and formally graded using the NOS system for observational, case-control and RCTs. Incidence rates were tabulated for the following groups: never infected (NI) and unvaccinated (UV), NI and vaccinated (V), previously infected (PI) and UV, PI and UV. Pooling was performed by grouping the RCTs and observational studies separately, and then all studies in total. Risk ratios and risk differences are reported for individual studies and pooled groups, in 1) NPI/V vs. PI/UV and 2) PI/UV vs. PI/V analysis. In addition, number needed to treat (NNT) analysis was performed for vaccination in naïve and previously infected cohorts.

**RESULTS:** Nine clinical studies were identified including three randomized controlled studies, four retrospective observational cohorts, one prospective observational cohort, and a case-control study. The NOS quality appraisals of these articles ranged from four to nine (out of nine stars). All of the included studies found at least statistical equivalence between the protection of full vaccination and natural immunity; and, three studies found superiority of natural immunity. Four observational studies found a statistically significant incremental benefit to vaccination in the COVID-recovered individuals. In total pooled analysis, incidence in NPI/V trended higher than PI/UV groups (RR=1.86 [95%CI 0.77-4.51], P=0.17). Vaccination in COVID-recovered individuals provided modest protection from reinfection (RR=1.82 [95%CI 1.21-2.73], P=0.004), but the absolute risk difference was extremely small (AR= 0.004 person-years [95% CI 0.001-0.007], P=0.02). The NNT to prevent one annual case of infection in COVID-recovered patients was 218, compared to 6.5 in COVID-naïve patients, representing a 33.5-fold difference in benefit between the two populations.

**CONCLUSIONS:** While vaccinations are highly effective at protecting against infection and severe COVID-19 disease, our review demonstrates that natural immunity in COVID-recovered individuals is, at least, equivalent to the protection afforded by full vaccination of COVID-naïve populations. There is a modest and incremental relative benefit to vaccination in COVID-recovered individuals; however, the net benefit is marginal on an absolute basis. COVID-recovered individuals represent a distinctly different benefit-risk calculus. Therefore, vaccination of COVID-recovered individuals should be subject to clinical equipoise and individual preference.

## INTRODUCTION

With the emergence of the COVID-19 pandemic beginning early 2020, the rapid development and release of effective COVID-19 vaccinations represents a crowning achievement of the pharmaceutical and medical establishment. In the United States, Pfizer/NBiotech and Moderna achieved Emergency Use Authorization (“EUA”) for the use of novel mRNA vaccines in general populations. Later, Johnson and Johnson (J&J) was awarded an EUA for a one-dose viral vector vaccination. The efficacy of these vaccines are excellent, with the Pfizer and Moderna vaccinations reported to achieve 90.3-97.6% [1] and 89.3-96.8% [2] efficacy, and the J&J viral vector vaccine to be in the range of 55-74% [3]. The overall risk of severe adverse effects is generally considered to be extremely low [1,2,3].

While COVID-19 vaccinations are generally recommended for all persons 12 years of age and older without contraindications [4], the risk/benefit calculus may differ for individuals who may not expect the same benefit, or may be at higher risk of adverse effects. One major subpopulation in this category are those individuals who were previously infected with SARS-CoV-2 and recovered (i.e., “COVID-Recovered”), which now exceed 180 million persons worldwide [5]. Significant public debate is now occurring as to whether recovered COVID-19 patients possess sufficient natural immunity, and if there is any substantial incremental benefit to COVID vaccination. In the United States, the Center for Disease Control (CDC) currently recommends vaccinations in previously infected individuals without exception.

From early in the start of the pandemic through November 2020, a growing number of case reports and small series were published demonstrating the possibility of reinfection in previously infected individuals [6], however, the incidence and risk factors were only hypothesized. In December of 2020, Abu Raddad et. al. published a study of a Qatari population of 133,266 previously infected individuals and found a positivity and symptomatic incidence of reinfection of 0.18% and 0.04%, respectively, over a 6 month period [7]. More recently, Lawandi et. al. reported a retrospective cohort study spanning 238 U.S. hospitals and 131,773 patients and found suspected reinfection in 0.2% of patients [8]. A systematic review of the literature in May 2021, found that SARS-CoV-2 reinfection was an uncommon event, ranging from 0-1.1%, with no study reporting an increase in infection risk over time [9].

Similarly, several investigators have reported longitudinal and observational studies, that directly compare the relative incidence of reinfection in COVID-recovered persons to COVID-naïve populations, and found at significant risk reduction via natural immunity. In January 2021, Hanrath et. al. reported a series of 17,126 health care workers (HCW) in the U.K., and found 0% risk of reinfection, compared to a 2.9% positivity rate, resulting in a 100% risk reduction due to prior infection (P<0.0001) [10]. In February of 2021, Lumley et. al. published an observational cohort study of 12,541 HCWs, with known seropositive or seronegative status, and found an adjusted risk reduction 89%, with no symptomatic infections in seropositive individuals [11]. Hall et. al. published the results of prospective study of 30,625 participants, and reported an 84% risk reduction, with a minimal median protective effect of at least 7 months after primary infection [12]. Vitale et. al. published a report on 15,075 individuals in the Italian general population, and found a 94% risk reduction lasting at least one year. Leidi et. al. studied an observational cohort of 10,547 Swiss essential workers, and also found a 94% risk reduction lasting at least 8 months [13]. A fewer number of studies reported more modest risk reduction, but greater than 74% [14-16].

With the phenomena of reinfection in the COVID-recovered considered to be relatively low, and with the protective effect of previous infection on par with the major available COVID-19 vaccinations, the next important question is what is the comparative benefit of vaccination in COVID-recovered individuals? In this systematic review, we focus on the existing clinical studies that comparatively delineate the efficacy of natural immunity in COVID-recovered persons, as well as the incremental benefit of vaccination in this same population. We also perform a pooled analysis of the eligible literature in order to aggregate and add power to the findings. As this is an evolving, yet substantially important question, we consider published and pre-published studies, and independently appraise the strength of findings.

This review has important policy implications, as vaccine mandates are emerging in the public and private sectors. With few exceptions, these mandates do not generally grant natural immunity persons the same status as COVID-naïve individuals who have been “fully vaccinated”. If, however, the evidence objectively shows equivalence in protection, then these civic policies of vaccination in the COVID-recovered should be seriously questioned on the basis of medical necessity, ethical principles and legal precepts governing the maintenance of bodily integrity.

## METHODS

### Search Strategy and Data Extraction

We performed a qualitative and inclusive systematic review guided by the principles outlined in the Preferred Reporting Items for Systematic Reviews and Meta-Analysis (PRISMA) guidelines published in 2020. We used PubMed as our primary database for published and peer-reviewed articles, and MedRxIV for pre-published studies. We also included publically available briefings or communications available on the FDA and CDC website, given their central position in the dynamics of the pandemic. These briefings included significant data on subgroups, that were not presented in the academic literature. The search period was defined as December 1, 2020 to August 31st, 2021. This period was selected to include vaccination efficacy trials that occurred before the large-scale administration of vaccinations. Search words included “COVID-19”, “SARS-CoV-2”, “Coronavirus”, in combination with terms, “previously infected”, “reinfection”, “recovered”, “convalescent”, “natural immunity”, “recurrence”, “antibody”, “seropositive”, intersecting with searches on “vaccination” or “immunization”.

Articles included were screened independently by the authors, by title, or if necessary the abstract or available manuscript. Our review was limited to original articles of clinical outcome studies published in the English language, without limitations on age, gender, geography of the study population, or vaccine manufacturer. Selected articles were inclusive of direct comparisons of unvaccinated COVID-recovered individuals (determined either by RT-PCR, antigen or serology), to other populations (either vaccinated or unvaccinated). We excluded case reports, other systemic or scoping reviews, opinion publications, guidelines, comments, editorials, animal studies, *in vitro* studies (typically testing for immune response outcomes), duplicate studies, or studies not electronically accessible. We also excluded studies in which study size was less than 500 total participants in the previously infected or baseline seropositive cohorts. All listings were reviewed, and either included or excluded by the authors.

### Quality Appraisal of Identified Studies

Of the identified studies, each study was appraised. Observational, case-control and randomized controlled studies were assessed according to the respective Newcastle-Ottawa Scales (NOS), segmented by selection, comparability and outcome quality. Criteria on these scales were applied consistently, across studies. Additionally, each study was qualitatively assessed with regards to study design, patient/case definitions, methodology, and outcome measurement

### Incidence Analysis and Study Pooling

Four groups were constructed: never previously infected and unvaccinated (NPI/UV), never previously infected and vaccinated (NPI/V), previously infected and unvaccinated (PI/UV), and previously infected and vaccinated (PI/V). In each of the identified studies, these cohorts were identified from the primary data, and event (infection) numbers, total cohort size, and time at risk (in person-years) was tabulated. While most data was available in the manuscripts and briefings, and some required personal communication with authors. If we did not get a reply from the authors, we made conservative assumptions (details in table) about the time at risk. Studies were excluded if it did not contain absolute event counts, or if the design of the study did not conform to a pool. All data was then tabulated to include the unadjusted infection events, total participants, and the person-time follow-up (converted to standardized person-years) for each study. We then conducted two fundamental comparisons. First, we analyzed the NPI/V versus PI/UV groups, to delineate the relative and absolute differences in protection between vaccination and natural immunity. Second, we compared the P/UV versus PI/V groups, to define the absolute and relative incidence differences due to vaccination within the PI group.

For each study and comparison, the risk ratio (RR) and absolute risk (AR) differences were calculated, along with 95% confidence intervals. A pooling method was performed combining the RCTs, the observational studies, and then all studies together, using Mantel-Haenszel (M-H) methods to arrive at pooled risk ratios and differences, along with confidence intervals and P values. M-H methods were utilized for its ability to provide pooled risk ratios across designated strata, in this case, randomized studies versus observational studies. A P-value of 0.05 was used as the threshold for significance. For the PI/UV vs PI/V analysis, the risk differences (1/AR) were used to generate a number needed to treat (NNT) per annum, for the NPI and PI pooled groups.

## RESULTS

### Search Outcome

We began with the known FDA Vaccine Related and Biological Product Advisory Committee (VRBPAC) briefings for Pfizer, Moderna, and J&J [17,18,19]. These VRBPAC briefings contained significant subgroup data that was not included in the academic publications [1,2,3]. From the initial database searches in PubMed, we identified 234 results of which three met criteria [22,23,24]. A similar search in MedRxIV identified 422 studies, three studies [20,21,25]. Therefore, we identified a total of nine clinical studies reporting rates of infection in COVID-recovered, COVID-naïve, vaccinated and unvaccinated individuals. Three of these studies were randomized controlled studies (RCTs) [17,18,19] with subgroup analysis. Four of these studies were classified as retrospective observational cohort studies [21,22,24,25]. One study was a prospective observational cohort [20] and another was a case-control study [23]. Figure 1 depicts the PRISMA flow diagram. Table 1 summarizes the identified studies, listed in chronological order of publication.

**Figure 1.**
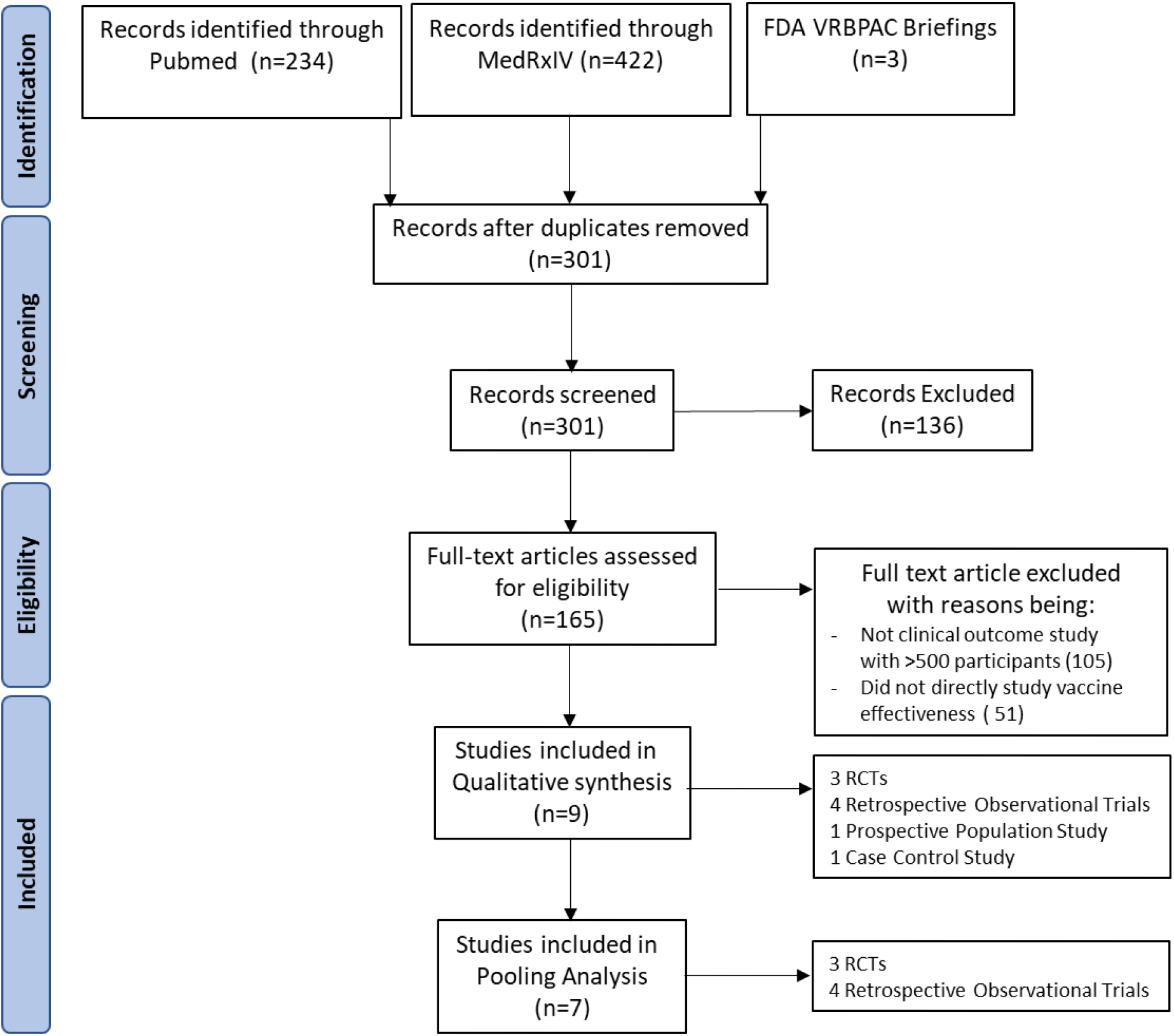
PRISMA flowchart

**Table 1.**
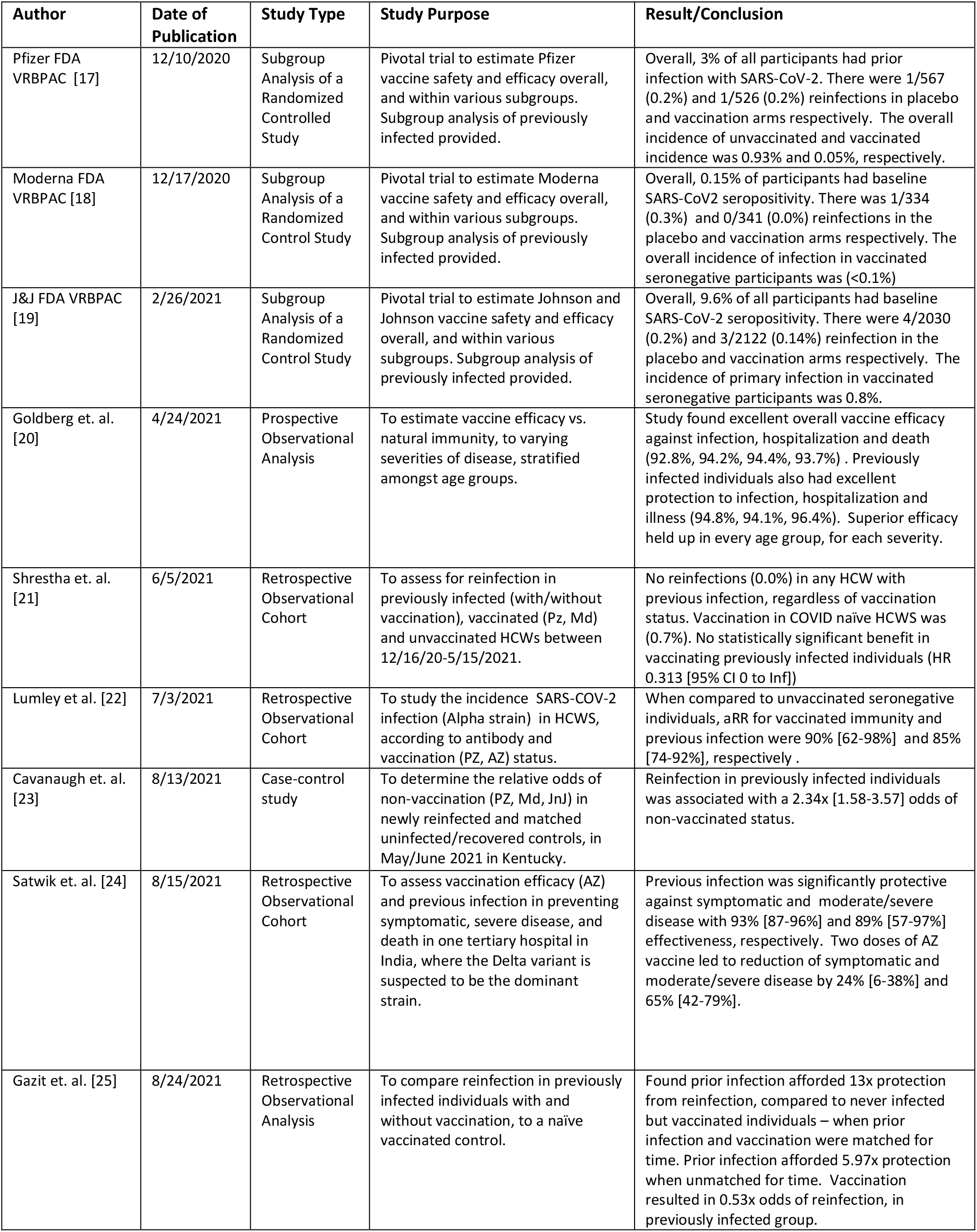
Description of identified clinical studies, type, purpose and conclusion.

### Description and Appraisal of Identified Studies

Each study was meticulously reviewed qualitatively, and objectively appraised based upon the respective NOS rating scales and other characteristics. Table 2 depicts the strengths and weaknesses of each study, and the NOS score.

**Table 2.**
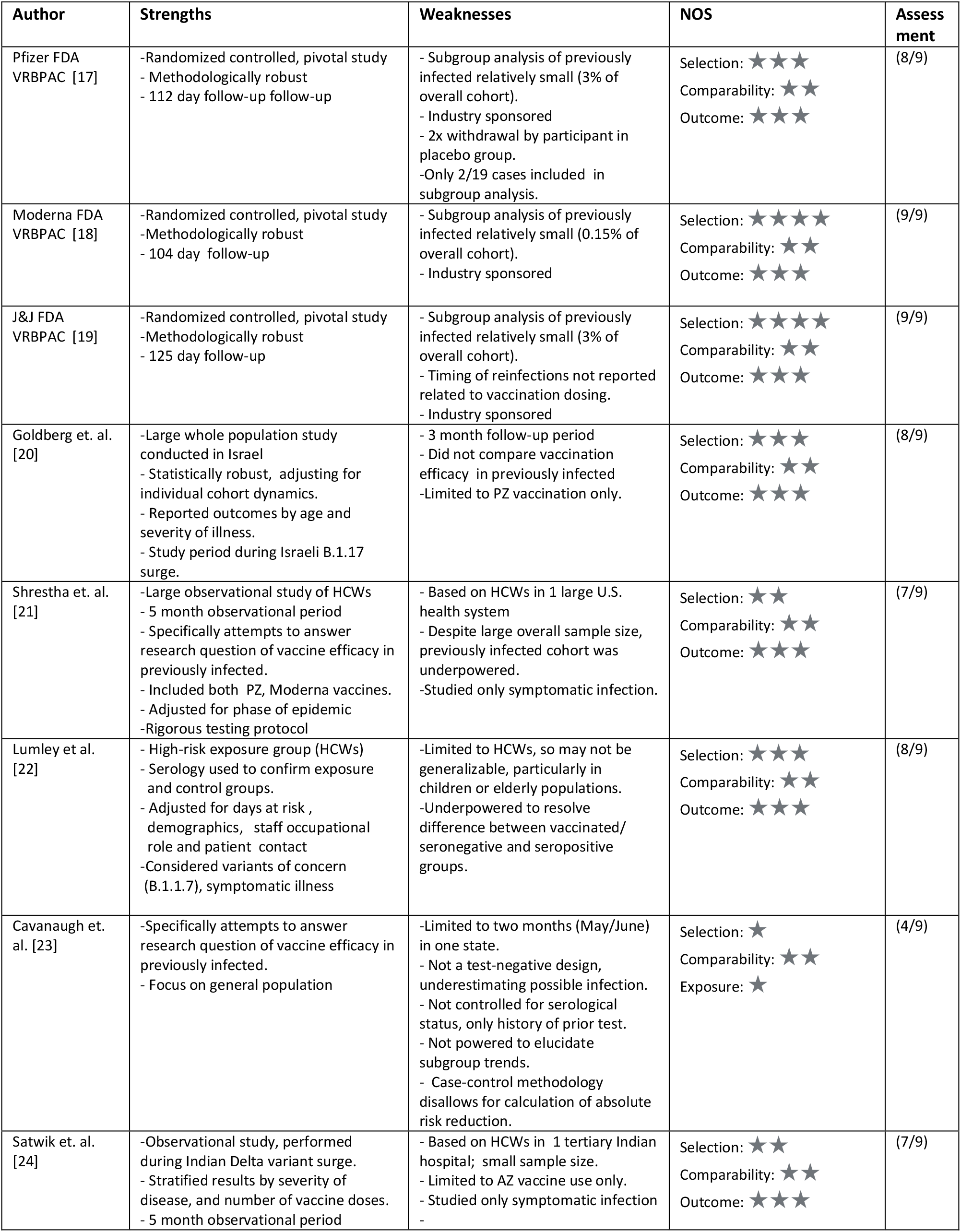
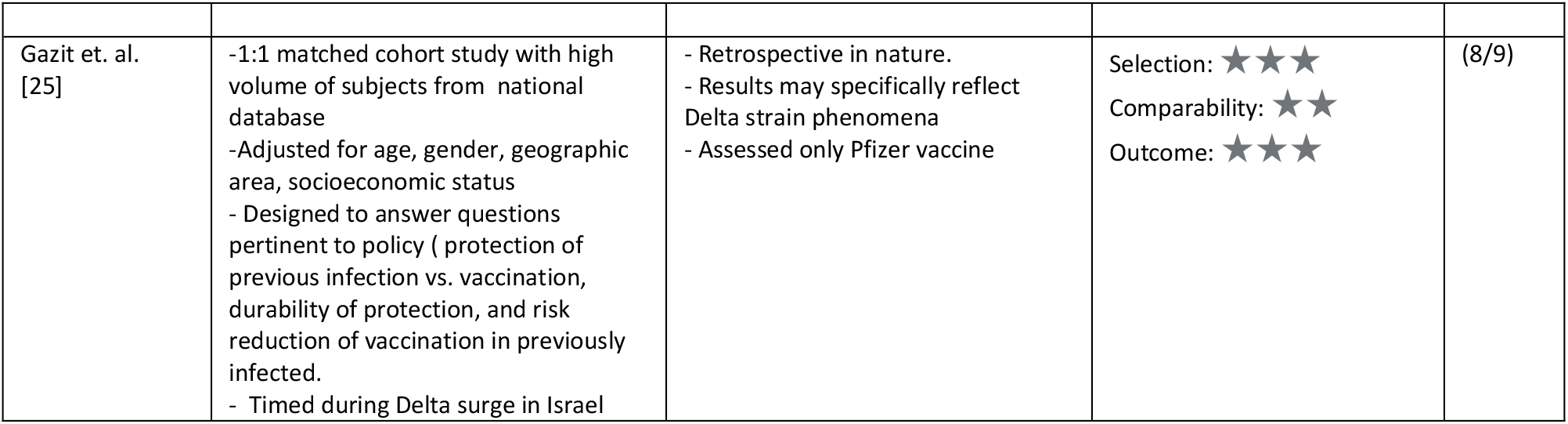
Strengths, weaknesses, and appraisals of identified studies.

The pivotal vaccine trials (Pfizer, Moderna, and J&J) each contained subgroups of participants that were previously infected, either by medical history or serological status. These studies were large and methodically regulated randomized controlled trials (RCTs), specifically designed to assess for vaccine efficacy in a general population. None of these three studies could statistically conclude that vaccination in the previously infected and/or serologically positive population would benefit from the vaccine, due largely to the small overall incidence of reinfection. In the Pfizer trial, when observing all participants from enrollment (without exclusion), there were a total of 19 reinfections (9 in placebo arm, and 10 in vaccination arm), however, the exact timings in relation to the second dose are not reported in the briefing. Overall, these are well-conducted trials, with the limitations of power for this subgroup analysis, and relatively short follow-up (3 to 4 months), with differing follow-up periods within various phases of the pandemic. Critically, the Moderna and J&J trials achieve a full score (9/9) for NOS evaluation. The Pfizer trial received an 8/9 score, with one star deducted due to lack of confirming the presence or absence of current or prior infection, prior to intervention.

Goldberg et. al. [20] released a study set in the unique situation of Israel, which undertook a massive vaccination campaign, but during the study period, previously infected individuals were explicitly excluded from vaccination. This policy allowed for a direct large volume and prospective comparison of COVID-naive vaccinated individuals to COVID-recovered unvaccinated individuals. The overall study population included 6.3 million individuals 18 years and older, and utilized a dynamic cohort model that accounted for individuals progression through first dose to full vaccination status. The statistical methodology was robust, executing a Poisson regression, and adjusting for age, gender, prior PCR results, and municipal risk. Overall, the results found excellent vaccine efficacy in the NPI/V group of 92.8%, 94.2%, 94.4% and 93.7% against infection, hospitalization, severe illness and death, respectively. However, protection in the PI/UV cohort was superior with 94.8%, 94.1%, 96.4% against infection, hospitalization and severe illness. There were so few deaths in the PI/UV cohort, that it could not be statistically calculated. The trend of superior protection from natural immunity held up in every age demographic, for all severities of illness. Additionally, this study was conducted during the Israeli surge of the B.1.1.7 (Alpha) variant, suggesting robust natural immunity to variants of concern. Limitations of this study include a bias towards symptomatic testing, and the lesser likelihood of vaccinated or previously infected individuals getting symptomatic testing. As such, incidence rates may be underestimated, but not for more severe illnesses. In terms of appraisal, the study achieves 8/9 stars, with only one star deduction for the inability to confirm current infection prior to vaccination.

Shrestha et. al. [21] performed an observational study in the context of occupational health, set at the Cleveland Clinic, OH, USA. A total of 52,238 employees were enrolled, of which 2,579 had a history of previous SARS-CoV-2 infection. Of these previously infected individuals, 53% remained unvaccinated during the course of the observation period. Throughout the entire study, not a single previously infected individual (0%) presented with reinfection, regardless of vaccination status (PI/V and PI/UV). Consequently, the effective risk reduction by previous infection was effectively 100%. Conversely, the NPI/V cohort had a breakthrough of 0.7%. As expected, the vast majority of study positives were in the NPI/UV cohort. Using a Cox proportional hazard methodology, the authors found a significantly lower risk by vaccination in COVID-naïve individuals, but no significant difference in COVID-recovered persons attributable to vaccination. Limitations of this study include its definition of previous infection, which may neglect asymptomatic infections. Additionally, constraining the study to HCWs limits the study’s generalizability to demographics not represented in this particular workforce. With regards to NOS scoring, the study achieved 7/9 stars, with two star deductions for its limited focus on HCWs, and absence of demonstration that infection was present at the start of the study.

Lumley et. al [22] represents a high-quality observational cohort study, performed at Oxford University Hospitals, that evaluated the incidence of SARS-CoV-2 reinfection in 13,109 HCWs, stratified by serological and vaccination (one and two doses) status. Of note, this study coincided with the B.1.1.7 surge (Alpha) in the United Kingdom. There were a total of 327 infections in the study group, with 326 infections occurring in the seronegative unvaccinated or partially vaccinated group, and only one reinfection in the seropositive group. There were no infections in the vaccinated, seronegative group. The authors calculated a 90% and 85% risk reduction for vaccination in seronegative and seropositives, respectively, without statistical difference [P=0.96]). Additionally, the authors conducted a study on viral loads in symptomatic infection, and found the pre-vaccination seropositive cohort had the lowest viral loads in infected persons across the study. The authors concluded that “Natural immunity resulting in detectable anti-spike antibodies and two-dose vaccine does both provide robust protection against SARS-CoV-2 infection, including the B.1.1.7 variant”.

The strengths of Lumley et. al. are its robust methodology, that utilized a Poisson regression model to estimate the incidence per day-at-risk, adjusted for month, age, gender, ethnicity, staff role and direct patient contact. The cohort was drawn from a well-defined population of serologically tested individual, with either a positive or negative status prior to the study period. While the study pertains to a narrow population of HCWs, that are underrepresent certain age and ethnicity demographics in general populations, the high-risk setting contributes to the confidence of the result. The study was somewhat limited in its uncontrolled approach to testing, as some cohorts may have different tendencies to get tested. Finally, while the study was powered to resolve differences between unvaccinated seronegatives and the other cohorts, it was underpowered to determine superiority between fully-vaccinated seronegative and unvaccinated seropositive individuals, due to low rates in both groups. As such, the study informs on comparability in protection, but non-superiority of either vaccinated or seropositive status. Per our appraisal, the study rated 8/9 stars, with the only deduction being for lack of demonstration that infection was not present at the start of the study.

Cavanaugh et. al. [23] presented a case-control study, in the state of Kentucky, United States. The study used a linked state infection and vaccination databases, reconciled by name and date of birth. The authors identified 246 total “case” reinfections in May and June, 2021, drawn from all Kentucky residents aged ≥18 years, with a positive SARS-CoV-2 test in 2020. Case-patients were then matched 1:2 to control (492 individuals) non-reinfected patients, based on sex, age, and date of initial positive test. Unvaccinated individuals accounted for 72.8% of case-patients, whereas only 57.7% of the controls were unvaccinated. This calculates to an adjusted odds ratio (OR) of 2.34 (95% CI 1.58-3.47). The authors suggest, that “among persons with previous SARS-CoV-2 infection, full vaccination provides additional protection against reinfection.”

While Cavanaugh et. al. was specifically designed to assess for superiority of vaccination versus non-vaccination in previously infected individuals, the study had several limitations to consider. First, the study represents a single-state experience drawing only 246 reinfected patients in May and June of 2021 (out of potentially 275,000 eligible [26]), based upon a database matching algorithm, by which inefficient matching (duplicate names, incomplete records) could lead to disproportionate selection bias in this small sample. Second, the control group was not confirmed “test-negative”, and vaccinated individuals (symptomatic or asymptomatic) may be less inclined to get tested. Consequently, the case and control groups are not matched according to their likelihood of getting tested, which is a critical confounder. Third, case matching was only performed on the basis of age, gender, and month of previous infection; however, there are a number of other salient parameters that should have been addressed. For example, race, socioeconomics, and geography are all variables that could impact getting vaccinated and/or getting tested. Fourth, only reinfections reported in May and June of 2021 were used to identify case subjects, even though vaccinations were made available beginning December 2020. In terms of NOS assessment for case-control studies, it was rated 4 of 9 stars, with three star deductions in subject selection, and two star deductions in assessment and follow-up of outcome.

Satwik et. al. [24] reported a small observational study, performed on HCWs at one tertiary hospital in New Dehli, India, where primarily the Astra-Zeneca (ChAdOx1 nCov-19) vaccination was available for 4,296 employees. The authors report an effectiveness of 93% [95% CI 87-96%] versus two does vaccination efficacy of 24% [95% CI 6-38%], for all symptomatic infections. For moderate to severe disease, the effectiveness of previous infection was 89% [95% CI 57 to 97] versus 65% [95% CI 42-79%] for two dose vaccination. There were no deaths in the previous infection or two dose cohort. This study is notable for setting during the B.1.617.2 (Delta) variant surge, experienced in India during this time. A separate study performed at this institution, at the same time, noted approximately a 50% penetration of the Delta variant [27]. The underwhelming vaccine efficacy observed in this study was in line with other studies pertaining to the Delta variant, during the same observation period [28]. The limitations of this study are its relatively small size within a group of HCWs, lack of adjustments for basic demographics, testing of symptomatic individuals only, and primary use of the ChAdOx1 nCov-19 vaccine, which differs from other studies in this review. The study also occurred during a difficult period of Delta strain emergence, and led to shortened average follow-up in vaccinated individuals. Nevertheless, the authors conclude that “[previous infection offered] higher protection than that offered by single or double dose vaccine.” NOS assessment attributed 7 of 9 stars to this study, due to lack of confirming presence or absence of infection at start of the study, and the short-duration of follow-up, particularly in vaccinated cohorts.

Gazit et al. [25] recently presented a retrospective observational study, with a matched cohort analysis, in Israel during the Delta surge. The authors defined three groups: never infected and two doses of vaccination (Pfizer), previously infected and never vaccinated, and previously infected and one dose of vaccination (Pfizer). These groups then underwent a matched cohort comparison, controlling for age, gender, and geographic area, and socioeconomic status. When comparing the vaccinated COVID-naive group with the unvaccinated COVID-recovered in a matched timing analysis, they found a 13.06 (95% CI 8.08-21.11, P<0.001) increased risk of infection in the vaccinated cohort. For symptomatic infections only, the risk increased to 27.02-fold [95%CI12.7-57.5]). When time matching was removed, there still was a 5.96 [95% CI 4.85-7.33, P<0.001] increased risk of infection in the vaccinated no prior infection group. Finally, they compared vaccination to non-vaccination in previously infected individuals, and found a 0.53-fold risk reduction (95%CI 0.3-0.92, P<0.05). However, the absolute risk reduction was only 0.1% (17 cases/14,029 subjects). Similarly, for symptomatic individuals risk was reduced 0.68-fold (95%CI 0.38-1.21) with an absolute risk reduction of 0.04%, without reaching statistical significance. The authors bluntly conclude, “This study demonstrated that natural immunity confers longer lasting and stronger protection against infection, symptomatic disease and hospitalization caused by the Delta variant of SARS-CoV-2, compared to the BNT162b2 two-dose vaccine-induced immunity … [the previously infected] given a single dose of the vaccine gained additional protection against the Delta variant.”

The Gazit et. al. study was designed to specifically answer pertinent clinical questions, with robust methodology and adjustments. The strength of the study is the size of the cohorts and its matched design, that allowed for multivariable adjustments. The limitations of the study include its applicability primarily to the Delta variant and Pfizer vaccine only. As the authors only reported total events without respect to time, there could be time-varying complicating factors that alter the result. In terms of NOS rating, it achieved 8/9 stars, with its only deduction being the lack of demonstration of infection at the start of the study, due to its population database methodology.

### Risk Analysis: Vaccination vs. Natural Immunity (NPI/V vs. PI/UV)

The infection events per person were tabulated for each study, and are presented in Table 3, along with the risk difference and risk ratios between NPI/V and PI/UV groups. Figure 2 depicts a forest plot of these results graphically for individual studies, RCTs, observational, and total pooled groups. Four studies [19,21,24,25] favored an increased risk ratio in the vaccinated NPI/V group (three statistically significant), versus three studies suggesting increased risk in the natural immunity PI/UV group [17,18,22] (none statistically significant). This observation was similar in the absolute risk difference analysis. The pooled RCT studies led to an overall RR 0.59 [95% CI 0.04-8.28, P=0.69], whereas the RR for pooled observational studies was 3.71 [95%CI 1.75-7.86; P=0.0006]. Overall, the total pooled RR was 1.86 [95% CI 0.77-4.51, P=0.17]. Goldberg et. al. [20] was excluded from this analysis as numeric data for events and cohort sizes were not reported, however, it supports the pooled findings in finding superiority of natural immunity over vaccination. Cavanaugh et. al. [23] was also excluded as it only included analysis of previously infected individuals. Consequently, there was no study that could conclude superiority of vaccination protection over natural immunity with statistical confidence, but observational studies endorsed an advantage for protection by natural immunity.

**Table 3.** Summary of Reported Infection/Reinfection Incidences (per person, person-years)

| Study | NPI/UV<br>Cases (N)<br>%<br>Person-Years<br>Events/P-Y | NPI/V<br>Cases (N)<br>%<br>Person-Years<br>Events/P-Y | PI/UV<br>Cases (N)<br>%<br>Person-Years<br>Events/P-Y | PI/V<br>Cases (N)<br>%<br>Person-Years<br>Events/P-Y | NPI/V vs. PI/UV<br>RR [95% CI]<sup>4</sup><br>AR [95%CI]<sup>4</sup> | PI/V vs PI/UV<br>RR [95% CI]<sup>4</sup><br>AR [95%CI]<sup>4</sup> |
| --- | --- | --- | --- | --- | --- | --- |
| Pfizer [17] | 164 (17,720)<br>0.93%<br>2,242<br>**0.073** | 8 (17,637)<br>0.05%<br>2,237<br>**0.004** | 1 (567)<br>0.18%<br>60<br>**0.017** | 1 (526)<br>0.19%<br>56<br>**0.018** | 0.21 [0.03-1.69]<br>-0.013 [-0.046-0.019] | 0.93 [0.06-14.57]<br>-0.001 [-0.049-0.046] |
| Moderna [18] | 90 (14,730)<br>0.63%<br>2,787<br>**0.032** | 6 (14,312)<br>0.04%<br>2,387<br>**0.002** | 1 (334)<br>0.30%<br>59<br>**0.017** | 0 (341)<br>0.0%<br>60<br>**0.0** | 0.15 [0.02-1.21]<br>-.014 [-0.47-0.019] | 3.05 [0.13-73.39]<br>0.017 [-0.029-0.063] |
| J&J [19] | 509 (19,544)<br>2.60%<br>3,089<br>**0.164** | 173 (19,514)<br>0.89%<br>3,114<br>**0.056** | 4 (2030)<br>0.20%<br>321<br>**0.012** | 3(2122)<br>0.14%<br>336<br>**0.009** | 4.46 [1.67-11.93]<br>0.043 [0.029-0.058] | 1.40 [0.31-6.19]<br>0.004 [-0.012-0.019] |
| **RCT Pooled** | 763 (51,634)<br>1.48%<br>8,118<br>**0.094** | 187 (52463)<br>0.36%<br>7,738<br>**0.024** | 6 (2931)<br>0.20%<br>439<br>**0.014** | 4 (2989)<br>0.13%<br>453<br>**0.009** | 0.59 [0.04-8.28]<sup>5</sup><br>0.007 [-0.037--0.051]<sup>5</sup> | 1.45 [0.43-4.85]<br>0.004 [-0.010-0.019] |
| Shrestha et.al<br>[21] | 2139 (15,317)<br>14.0%<br>3,511<sup>6</sup><br>**0.609** | 15 (28,836)<br>0.05%<br>5,846<sup>6</sup><br>**0.002** | 0 (1265)<br>0.0%<br>256<sup>6</sup><br>**0.0** | 0 (1265)<br>0.0%<br>247<sup>6</sup><br>**0.0** | 1.36 [0.08-22.71]<br>0.0026 [-0.003-0.0081] | Not estimable<br>0.000 [-0.008-0.008] |
| Lumley et al.<br>[22]<sup>1</sup> | 635 (10,513)<br>6.0%<br>6,231<br>**0.102** | 2 (940)<br>0.21%<br>107<br>**0.019** | 12 (1273)<br>0.94%<br>543<br>**0.022** | 1 (974)<br>0.10%<br>92<br>**0.011** | 0.85 [0.19-3.73]<br>-0.003 [-0.032-0.025] | 2.03 [0.27-15.42]<br>0.011 [-0.013-0.036] |
| Satwik et. al.<br>[24]<sup>2</sup> | 128 (790)<br>16.2%<br>182<br>**0.685<sup>7</sup>** | 323 (2120)<br>15.2%<br>430<br>**0.696<sup>7</sup>** | 5 (147)<br>3.4%<br>36<br>**0.138** | 4 (596)<br>0.67%<br>129<br>**0.031** | 5.01 [2.21-11.32]<br>0.55 [0.4354-0.6775] | 4.48 [1.27-15.82]<br>0.108 [-0.0090-0.225] |
| Gazit et. al.<br>[25]<sup>3</sup> | - | 640 (46,035)<br>1.39%<br>9,459<br>**0.068** | 108 (46,035)<br>0.23%<br>9,459<br>**0.011** | 20 (14,029)<br>0.14%<br>2,882<br>**0.007** | 5.93 [4.84-7.25]<br>0.056 [0.051-0.062] | 1.65 [1.02-2.65]<br>0.005 [0.001-0.008] |
| **OBS Pooled** | 2902(26,620)<br>10.9%<br>9,925<br>**0.292** | 980(77,931)<br>1.26%<br>15,842<br>**0.062** | 125(48,720)<br>0.26%<br>10,296<br>**0.012** | 25 (16,819)<br>0.15%<br>3,352<br>**0.007** | 3.71 [1.75-7.86]<sup>5</sup><br>0.092 [0.031-0.153]<sup>5</sup> | 1.94 [1.17-3.21]<sup>5</sup><br>0.0038 [-0.0036-0.011]<sup>5</sup> |
| **Total Pooled** | 3665(78,254)<br>4.68%<br>18,042<br>**0.203** | 1167 (129,394)<br>0.90%<br>23,580<br>**0.049** | 131(51,651)<br>0.25%<br>10,735<br>**0.012** | 29(19,808)<br>0.15%<br>3,804<br>**0.007** | 1.86 [0.77-4.51]<sup>5</sup><br>0.049 [0.0084-0.0893]<sup>5</sup> | 1.82 [1.21-2.73]<sup>5</sup><br>0.0039 [0.001-0.007]<sup>5</sup> |

<sup>1</sup> Lumley et. al. included 1- and 2-dose vaccination groups. For this analysis, only the 2-dose vaccination group was counted.
<sup>2</sup> Satwik et. al. data not entirely provided in publication. Data obtained via personal communication.
<sup>3</sup> Gazit et. al. described three models of study, with a matching comparison. For this analysis, Model 2 was used for NPI/V, and PI/UV analysis, and Model 3 was used for the PI/V group. An NPI/UV group was not reported in this study, and therefore omitted from analysis.
<sup>4</sup> Risk ratios and differences calculated from Events/Person-Year figures
<sup>5</sup> Pooled RR and risk differences adjusted and determined by M-H methods.
<sup>6</sup> Time-at-risk not published in Shrestha et. al. manuscript. Estimated by taking number of people in group, multiplied by length of study (in years)
<sup>7</sup> Noted that there is no difference in incidence between NPI/UV and NPI/V groups. The authors of this study were contacted, and relayed at vaccine efficacy of 28%, after adjustment by a Cox proportion regression. Additionally, the author notes the implementation of vaccination during the peak phase of Delta, and the shorter follow-up in the vaccinated cohort.

**Figure 2.**
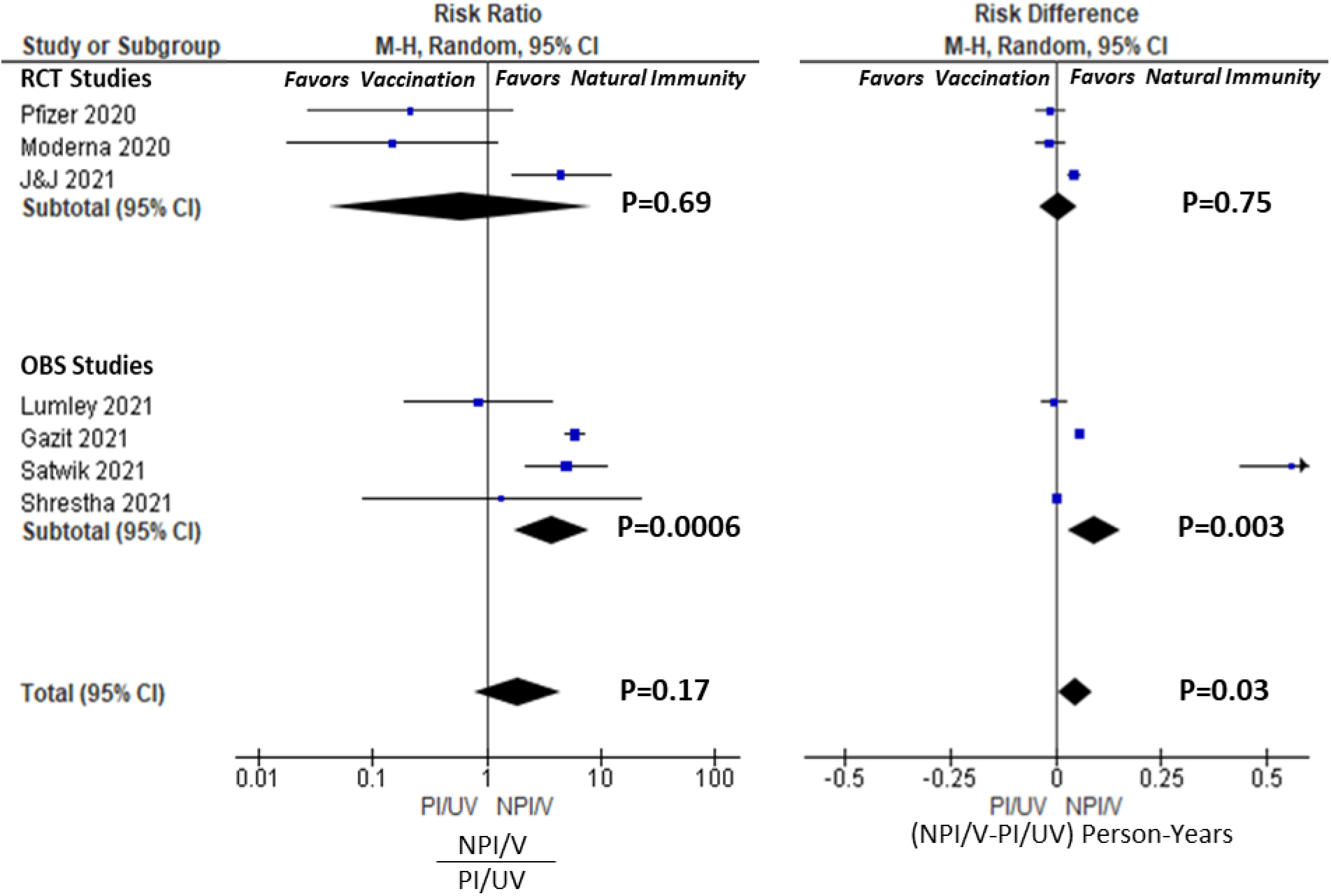
Forest plots for NPI/V vs. PI/UV Analysis. Risk ratio and differences calculated for individual studies, and RCT, OBS and Total pools. Risk ratio calculated as incidence ratio of (NPI/V)/)PI/UV). Risk difference calculated incidence difference of NPI/V-PI/UV.

**Figure 3.**
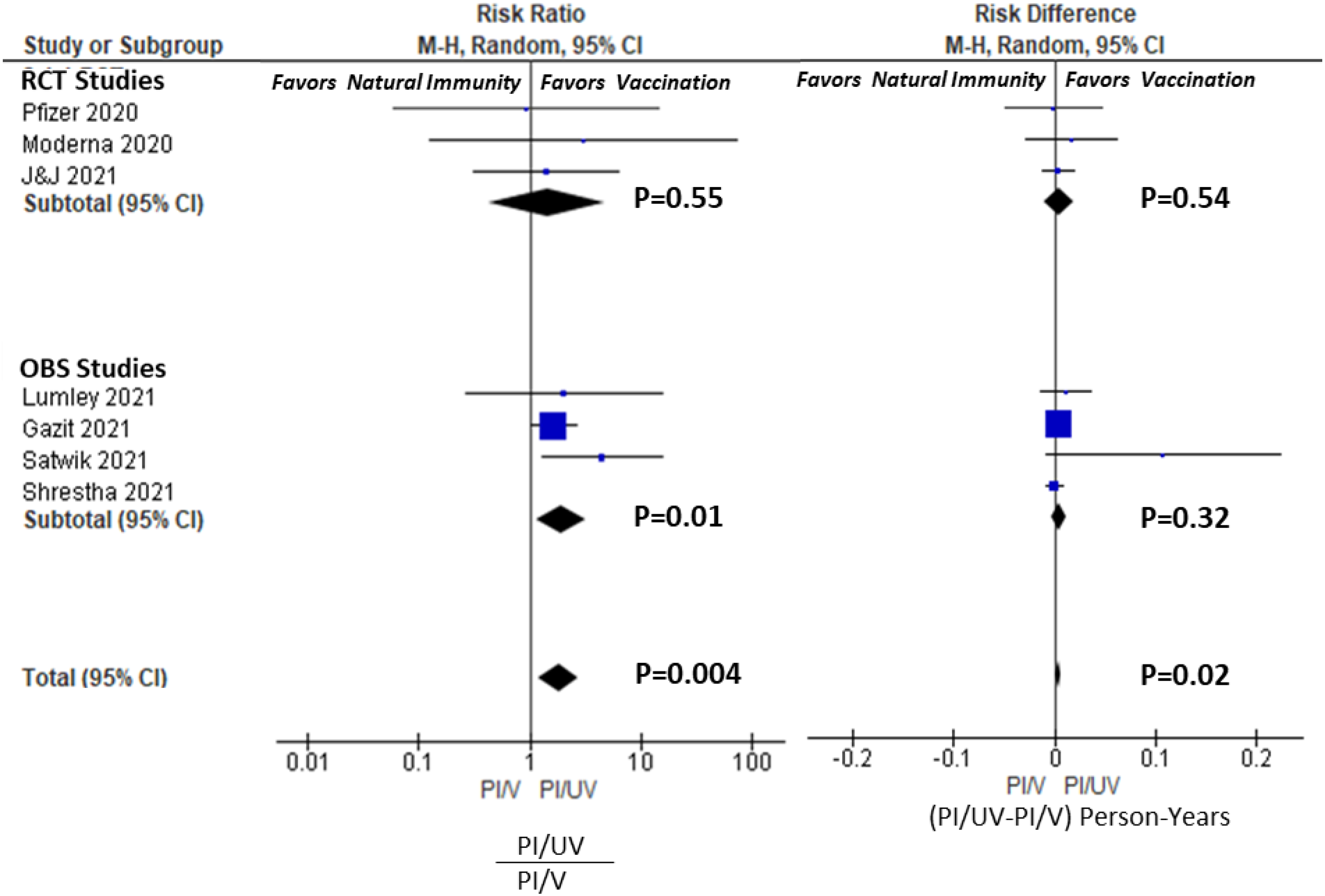
Forest plots for PI/UV vs. PI/V Analysis. Risk ratio and differences for individual studies, RCT, OBS and Total pools. Risk ratio calculated as incidence ratio of (PI/UV)/(PI/V). Risk difference calculated as incidence differences of PI/UV-PI/V.

### Risk Analysis: Vaccination vs Non-vaccination in the Previously Infected (PI/V vs. PI/UV)

Table 3 also summarizes the incidence rates, the RR and AR for reinfection in the PI/V vs. PI/UV groups. None of the individual RCTs found a statistically significant advantage to vaccination in preventing reinfection, and the RCT pooled results resulted in a RR of 1.45 [95%CI 0.43-4.85, P=0.55], and the AR of 0.004 person-years [95%CI -0.010-0.019, P=0.54]. However, three of the four observational trials did find an advantage to vaccination in the previously infected with an RR of 1.94 [1.17-3.21, P=0.01], but the AR was only 0.004 person-years [95%CI -0.004-0.011, P=0.32]. Overall, in the total pooled group, there was significant protection to vaccination in COVID-recovered persons, with an RR of 1.82 [1.21-2.73, P=.004], but this effect was modest on an absolute scale, 0.004 [95%CI 0.001-0.007,P=0.02]. Table 4 displays the applicable NNTs for each RCT, observational and total pools. The NNT by vaccination in COVID-recovered individuals in the 218, 207, and 214 persons in Total, RCT, and Observational pools, respectively. Comparatively, the NNTs in COVID-naïve individuals were 6.5, 14.3, and 4.3 persons, respectively. In the Total pool, this represents 33.5-fold larger population that needs to be vaccinated, in order to prevent one case of COVID per year.

**Table 4.**
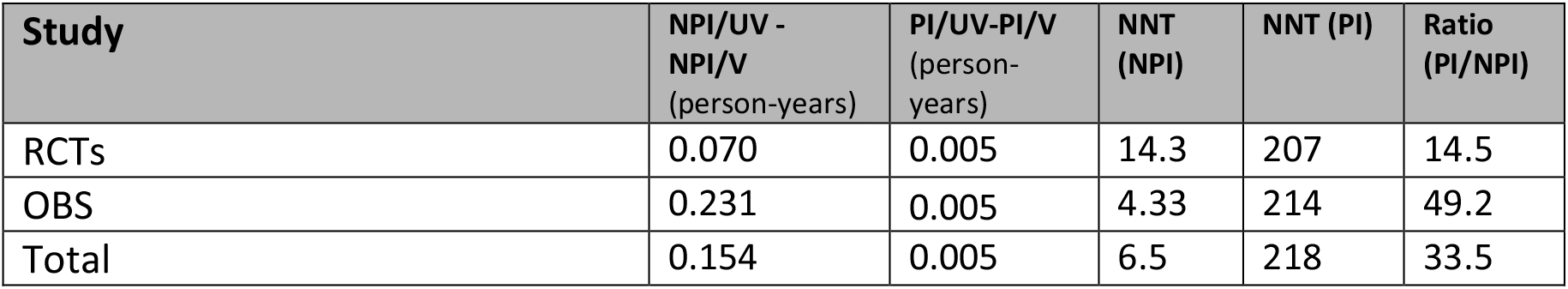
Summary of Vaccination Risk Difference and NNT in Pooled Populations.

## DISCUSSION

Despite the massive investigational attention on SARS-CoV-2 infection, COVID illnesses and vaccine efficacy, our systematic review identified only a relatively few outcome studies that investigated the comparative benefit of vaccination in COVID-recovered individuals. There are two relevant but separate questions that need to be considered. First, does previous infection protect individuals to the same degree as what we currently consider “full vaccination” in COVID-naïve individuals? Second, is there an incremental benefit of vaccination to previously infected individuals? Table 5 summarizes the significance of our findings, and each question is considered below. In summary, our analysis demonstrates that natural immunity in the COVID-recovered performs better than full-vaccination alone in COVID-naïve persons, however, there is a small absolute benefit to vaccination in COVID-recovered persons.

**Table 5.** Summary of Pooled Analysis Results and Statistical Significance

| Pool | NPI/V vs. PI/UV<br>RR<br>AR | PI/UV vs. PI/V<br>RR<br>AR |
| --- | --- | --- |
| RCTs | P>0.05<br>P>0.05 | P>0.05<br>P>0.05 |
| OBS | Favors Natural Immunity<br>Favors Natural Immunity | Favors Vaccination<br>P>0.05 |
| Total | P>0.05<br>Favors Natural Immunity | Favors Vaccination<br>Favors Vaccination |

### Does natural immunity provide at least equivalent protection from infection to that afforded by vaccination in the COVID-naïve persons?

The CDC currently recommends vaccination in all individuals 12 and older, regardless of a history of previous infection, under the assumption that recovered individuals are still at risk for reinfection and transmission. However, as discussed earlier, the data indicate that this reinfection rate is low. From a policy perspective, it is relevant to understand if natural immunity in COVID-recovered individuals provides similar protection from reinfection compared to vaccination in COVID-naïve persons, given the newfound social status of being “fully vaccinated”. We emphasize that it is highly unadvisable for COVID-naïve persons to seek infection as a means to avoid vaccination, and the risk of COVID illness (serious or otherwise) far exceeds the risk of vaccination. However, if natural immunity is at least equivalent, to some brands of vaccination (i.e., adenoviral vs. mRNA COVID-19 vaccines), then any rigid mandate to vaccinate COVID-recovered individuals would be of questionable legal and ethical standing, on the basis of suspect medical necessity and even a potential for harm.

In our systematic review, there was no clear evidence that vaccinated COVID-naïve individuals enjoyed greater protection than unvaccinated COVID-recovered counterparts. In fact, four observational studies [20,24,25] and one RCT [19] found that natural immunity provided superior protection to vaccination in the COVID-naive. None of the identified studies, or pooled groups, found a statistically significant advantage to vaccination in the COVID-naïve population, although the RCTs trended in favor of vaccination. There was solid internal consistency between the conclusions of our pooled analysis, and the conclusions of the authors of the studies, taken individually.

While RCTs are more methodologically sound, observational studies yield a more practically realizable result. In our study, pooled RCTs trended to favor vaccination, while the pooled observational studies more strongly supported natural immunity. This disparity can be partially explained by the difference in timing of the studies, as the RCTs were performed in an earlier phase of the pandemic compared to the observational trials which occurred during the emergence of variants. For example, Satwik et. al. (India) and Gazit et. al. (Israel), which occurred during the mid-2021 Delta phases of the pandemic, found stronger and statistically significant benefits of natural immunity compared to vaccination alone. Gazit et. al. found 13-fold increased odds of all infection in the vaccinated COVID-naive compared to unvaccinated COVID-recovered individuals, and a 27-fold increase in symptomatic infection. These more recent and stronger findings of stronger natural immunity highlight the time-dependent sensitivity of this analysis, due to variants of concern and potential waning vaccination efficacy. This observation is in line with other recent studies by Keehner et. al. [29] and Goldberg et. al. [30], that demonstrated a time-dependent decline in vaccine efficacy during the Delta phase of the pandemic..

In total, the evidence points quite convincingly to at least the equivalency between the protection of natural versus vaccinated immunity, with the possibility of enhanced durability of protection from natural immunity in non-controlled settings and later phases of the pandemic.

### Does vaccination in the previously infected provide a reduction in risk of reinfection ?

With regards to this question, the vaccination RCTs did not find a significant benefit to vaccination in the previously infected, either individually or in pooled RCT analysis. This conclusion is reflected in the official briefing narratives, which explicitly stated an absence of observed benefit of vaccination in recovered individuals, due primarily to the limitation of study power. In the pooled observational trials, however, the stronger relative effect was seen favoring vaccination, but the absolute effect was still small. Overall, the total pooled results demonstrated a statistically significant 1.86x enhanced protection by vaccination in COVID-recovered persons, which generally agrees with Gazit et al. (1/.53=1.89x) and Cavanaugh et. al. (2.34x), the latter of which was not included in the pooled analysis. Generally, we can conclude that vaccination in the COVID-recovered roughly halves the risk of reinfection, based on our pooled results and individual studies.

However, on an absolute basis, the risk reduction is quite modest. This is most tangibly seen in our pooled NNT analysis, where 218 recovered individuals would need to be vaccinated in order to prevent one case of COVID annually, compared to only 6.5 COVID-naïve individuals. This represents a 33.5-fold difference the absolute effect size between COVID-naïve and COVID-recovered individuals.

This disparity in NNT highlights the muted absolute benefit of vaccination to COVID-recovered individuals, compared to that enjoyed by COVID-naïve individuals. While our systematic review did not specifically cover risk of vaccination, recent studies have shown that vaccinations have a small but excess risk of adverse events appears in the range of 2-80 events per 100,000 [31]. There are also some reports, though no consensus, that previously infected individuals may have an increased risk of local and systemic adverse effects [32]. Therefore, while vaccination is overwhelmingly safe for the general population, and even for most COVID-recovered individuals, higher-risk subgroups therein, are subject to a distinctly different risk/benefit calculus and narrower therapeutic window, suggesting that individual factors with clinical equipoise should be utilized. Further evaluation of adverse events specifically within COVID-recovered individuals is warranted, as is a formal evaluation of the risk/benefit calculus. Civil policies, including vaccine mandates, should strongly consider automatic exemption from vaccination based on history of prior infection or serological evidence of immunity, until the risk/benefit is better delineated.

### Strengths and Limitations

This systematic review has several strengths. First, it uses the available literature to address timely questions with relation to vaccination policy. Second, by using an inclusive criteria, including the use of FDA briefings and the MedRxIv pre-print server, it aggregated all possible evidence on the subject current to the time, in a rapidly evolving setting. Our systematic review standardizes the metrics of vaccine and natural immunity, by reporting both relative and absolute measures, in order to provide a broader perspective on policy implications. Our strategy of stratifying the pooling of RCTs, observational studies, and then in total pools, via M-H methods, also allows for the assessment of internal consistency of our conclusions.

However, this systematic review has its own limitations. First and foremost, the question of vaccine efficacy in recovered COVID individuals is rapidly evolving, with new data and studies continuously being published. Indeed, several of the studies included in this review are still in peer-review [20,21, 25], though we have objectively attempted to appraise the quality of each of these studies with use of the established NOS rating scale. Second, we included results from PubMed, MedRxiv, and FDA/CDC briefings, while excluding briefings from other international regulatory organizations and other pubic databases. This could lead to a selection bias in identified studies. Third, the comparability of these studies are not precise, with some variation in case and endpoint definitions, follow-up periods, pandemic phase, and other methodological variations (incidence calculation, statistical tests etc.). This can confound the direct comparison of results in between studies, and assumptions for pooling.

Finally, when considering pooling analyses, the question of study homogeneity needs to be considered. While the RCTs were relatively homogenous in their study design and methodology, the other observational and case-control studies had significant differences that warrant consideration. For example, Shrestha et. al., Lumley et al., and Satwik et al. considered only HCWs in single health-systems, whereas Goldberg et. al. and Gazit et al. considered entire national populations. Satwik et. al., appears to be somewhat of an outlier in the magnitude of its results (but not the trend of its conclusion), due to the rapid rise of Delta during the implementation of vaccination, and the short follow-up in this group. This is a different scenario than what is presented in Gazit et. al, where a significant portion of the Israeli population was already fully-vaccinated, during the study period. Other difference among studies was the difference in vaccination brand utilization, vaccination cadence (one versus two doses, and interval), definitions of endpoints, inclusion and exclusion criteria, timing within the phases of the pandemic, and variants of concern. Despite these disparities, the observations we have made are remarkably consistent across the included studies and pools: that protection of vaccinated and natural immunity are at least roughly equivalent; and, while there may be some incremental protection to vaccination in COVID-recovered individuals, the absolute magnitude of that protection is dramatically lower compared to that experienced by COVID-naïve individuals.

## CONCLUSION

Overall, our comprehensive systematic review identified nine clinical studies of various design, of which seven could be included in a pooled analysis. From review of these studies, we conclude that there is no statistical advantage to vaccination in the COVID-naive, compared to natural immunity in the COVID-recovered. Vaccination in the COVID-recovered may provide some incremental protective benefit, but the size of this benefit is marginal. Explicitly, COVID-naïve individuals should not seek infection to bypass vaccination. However, COVID-recovered individuals should be considered to have at least equal protection to their vaccinated COVID-naïve counterparts. The COVID-recovered also represent a unique population segment with distinct risk/benefit considerations and a narrower therapeutic window. National policy should reflect the need for clinical equipoise and restraint in the decision to vaccinate these individuals by mandate.

## Data Availability

All data is contained within the manuscript, and were obtained from included references or personal communication.

## ACKNOWLEDGEMENTS

The authors would like to acknowledge authors of the included studies, who provided advice and unpublished data required to complete this study.

